# Dissecting Disease Tolerance in *Plasmodium vivax* Malaria Using the Systemic Degree of Inflammatory Perturbation

**DOI:** 10.1101/2021.03.06.21253036

**Authors:** Caian L. Vinhaes, Thomas A. Carmo, Artur T. L. Queiroz, Kiyoshi F. Fukutani, María B Arriaga, Marcus V. G. Lacerda, Manoel Barral-Netto, Bruno B. Andrade

## Abstract

Homeostatic perturbation caused by infection fosters two major defense stratagems, resistance and tolerance, which promote the host’s survival. Resistance relates to the ability of the host to restrict the pathogen load. Tolerance minimizes collateral tissue damage without directly affecting pathogen fitness. These concepts have been explored mechanistically in murine models of malaria but only superficially in human disease. Indeed, individuals infected with *Plasmodium vivax* may present with asymptomatic malaria, only mild symptoms, or be severely ill. We and others have reported a diverse repertoire of immunopathological events that potentially underly susceptibility to disease severity in vivax malaria. Nevertheless, the combined epidemiologic, clinical, parasitological, and immunologic features associated with defining the disease outcomes are still not fully understood. In the present study, we perform an extensive outlining of cytokines and inflammatory proteins in plasma samples from a cohort of individuals from the Brazilian Amazon infected with *P. vivax* and presenting with asymptomatic (n=108) or symptomatic (n=134) disease (106 with mild presentation and 28 with severe malaria), as well as from uninfected endemic controls (n=128) to elucidate these gaps further. We employ highly multidimensional Systems Immunology analyses using the molecular degree of perturbation to reveal nuances of a unique profile of systemic inflammation and imbalanced immune activation directly linked to disease severity as well as with other clinical and epidemiologic characteristics. The findings mapped the relationships between the systemic degree of inflammatory perturbation and parasitemia values to define the disease tolerance in vivax malaria.

**Author Summary:** *Plasmodium vivax* infection can result in a broad spectrum of disease manifestations, ranging from asymptomatic malaria to severe life-threatening disease. Despite significant advances in the current understanding of the critical factors associated with the disease outcomes in vivax malaria, the immunopathological events responsible for the diversity of severe manifestations in the disease remain deeply unknown. Here, a large panel of cytokines/chemokines were assessed in plasma samples from a Brazilian cohort of *P. vivax* patients presenting with asymptomatic infection or symptomatic malaria at the time of diagnosis, as well as from uninfected endemic controls, to define the relationships between systemic inflammation, disease presentation, parasitemia, and epidemiologic characteristics. In-depth analyses using the molecular degree of perturbation were employed to reveal nuances of a unique profile of systemic inflammation and imbalanced immune activation directly linked to disease severity. Moreover, the discoveries diagrammed the occurrence of disease tolerance by narrowing down the interactions between the systemic degree of inflammatory perturbation and parasitemia values in vivax malaria patients.

## Introduction

Malaria remains a major global cause of morbidity and mortality, with devasting impact over the last 10 years, estimated by more than 200 million cases and 400,000 deaths reported per year worldwide [1]. In contrast to the burden of *Plasmodium falciparum* infection in African countries, *Plasmodium vivax* represents the predominant parasite in American regions, with noticeable incidence within the Brazilian Amazon, responsible for the vast majority of malaria cases in this area [1, 2]. Also, infection with *P. vivax* is historically thought to result in milder disease presentation than that caused by *P. falciparum*, mostly because the latter commonly results in high parasitemia, intense hemolysis and inflammation associated with cytoadherence to small capillaries (reviewed in [3]). Nevertheless, more recently, several clinical and immunologic reports have revealed that *P. vivax* infection can clearly result in severe clinical presentations such as respiratory distress and acute kidney failure [4–6]. The exact immune mechanisms that underly susceptibility to infection and disease severity upon infection with *P. vivax* are not yet fully understood. We and others have described that older individuals who live for many years in a highly endemic area, and those who are highly exposed and had several previous malaria episodes, tend to develop asymptomatic *P. vivax* infection, which is associated with very low parasitemia and diminished inflammatory responses [7]. Furthermore, a broad spectrum of disease manifestations may occur among those who get sick, ranging from oligosymptomatic condition to multi-organ damage [8]. Understanding the molecular determinants of disease severity may help develop improved prophylactic and therapeutic strategies and optimize patient care.

The capacity to restrict the infection burden and control inflammation and immune activation are major determinants of the malaria clinical outcomes (reviewed in [9, 10]). The ability to control parasite load results from anti-microbial immune responses and leads to pathogen elimination and resistance to infection [11]. Exacerbated anti-parasite immune responses promote oxidative stress and inflammatory burst, which in turn cause collateral tissue injury, named immunopathology [12]. In this scenario, the capability to minimize organic damage without directly influencing the infection burden or pathogen fitness is a critical survival strategy defined as disease tolerance [10,11,13–15]. In the context of malaria, disease tolerance is present in highly exposed individuals from endemic regions, where subjects may harbor a wide range of detectable parasitemia levels without exhibiting severe clinical manifestations [16–18]. Findings from previous investigations support the idea that disease tolerance in malaria may occur as modulation of pro-inflammatory responses and dendritic cell function [19, 20].

Furthermore, in experimental malaria models, control of the collateral tissue damage occurs through tightly regulated mechanisms such as glucose and iron metabolism, renal control, and adrenal hormones [15,21–23]. In humans, we have previously reported that metabolic adaptation to iron overload through induction of ferritin confers tolerance to *P. vivax* malaria [15]. Despite the substantial advance in the knowledge about the relevant determinants of tolerance to malaria, especially in murine models, to our knowledge no detailed characterization of the systemic inflammation and immune activation profiles related to this host defense strategy has been reported in *P. vivax* malaria. Here, we aimed to analyze the degree of systemic inflammatory imbalance in *P. vivax* patients from the Brazilian Amazon. We have compared individuals with asymptomatic, mild, or severe symptomatic disease, to define the complexity and multifactorial regulation subjacent to the malaria immunopathogenesis and the responses underlying the disease tolerance. To do that, we employed a recently reported approach named the molecular degree of inflammatory perturbation [24–28] using concentrations of a large panel of cytokines, chemokines, growth factors and acute phase proteins in peripheral blood at the time of malaria diagnosis. The findings from our in-depth analyses support the idea that disease tolerance in *P. vivax* malaria is a tightly regulated process that integrates features highly influenced by age, previous malaria episodes, and a coordinated regulation of the systemic concentrations of mediators of inflammation.

## Materials and Methods

### Ethics Statements

Written informed consent was obtained from all participants or their legally responsible guardians, and all clinical investigations were conducted according to the principles expressed in the Declaration of Helsinki. The project was approved by the institutional review board of the *Faculdade de Medicina*, *Faculdade São Lucas*, Rondônia, Brazil, where the study was performed (protocol no. AP/CEP/55/07).

### Study Design

We performed multidimensional retrospective analysis of a databank containing clinical, epidemiological and immunological data from 530 individuals from the Brazilian Amazon (Rondônia, Brazil) recruited between 2006 and 2007, as part of a project aimed at describing determinants of susceptibility to vivax malaria that was finalized in 2010. In this project, both active and passive malaria case detection were performed. These included home visits in areas of high transmission (active case detection), and study of individuals seeking care at the diagnostic centers of Brazilian National Foundation of Health (FUNASA) or in a municipal hospital in Buritis, Rondônia, Brazil (passive case detection) [6,29,35,39,50–53]. Diagnosis, recruitment details and clinical definitions were previous published [15,29,35,50–54]. To direct test the effects of *P. vivax* infection in the molecular degree of perturbation only patients with *P. vivax* monoinfection (n=242, from those 108 were classified as asymptomatic and 134 symptomatic participants) and healthy controls (n=128) were included. The exclusion criteria for the present study were: *P. falciparum* infection documented by both microscopy and nested polymerase chain reaction (PCR), HIV-1, hepatitis B and C infection, yellow fever, leptospirosis, tuberculosis, cutaneous leishmaniasis, diabetes mellitus, allergic diseases as asthma, cancer, chronic degenerative diseases, sickle cell trait and the use of hepatotoxic or immunosuppressant drugs. Blood collection for nested polymerase chain reaction (PCR) and other measurements (cytokines, chemokynes, hepatic enzymes, creatinine, fibrinogen, bilirubin levels, free heme and haptoglobin) were performed at the time of study enrollment, meaning that specimens were collected at diagnosis, during acute phase of disease in symptomatic patients, before treatment initiation. Individuals without symptoms were actively recruited in their residencies by active search, mostly in remote riverine communities and had thick blood smear samples and a small aliquot of blood in EDTA tubes collected for diagnostic screening by nested PCR. The diagnosis in those individuals was performed after the clinical visit and if a positive test for malaria was found, a second visit within 30 days was performed in order to search for appearance of malaria symptoms. A new sample collection was made, and a second round of diagnostic tests was performed. Subjects that remained with positive nested PCR for Plasmodium within this period of 30 days and presented no malaria-related symptoms, such as fever (axillary temperature >37.8°C), chills, sweats, myalgia, arthralgia, strong headaches, nausea, vomiting, jaundice, and severe asthenia during this period were considered as asymptomatic malaria cases. During the first clinical visit (previously to the results of the diagnostic tests) all the asymptomatic individuals received counselling and were oriented to seek for health care in a malaria reference center in case symptoms appeared during the period between the 2 clinical visits. After the second clinical visit, all the patients that had positive malaria screening with microscopy and/or nested PCR were treated following the treatment guidelines of the Brazilian Ministry of Health. The plasma samples used to assess the biomarkers were collected during the second clinical visit, before the initiation of anti-malarial drugs, as previous described [39]. Clinical, demographic and epidemiological characteristics of the study participants were included in S1 Table. Considering the absence of a consensus to define severe vivax malaria, here we used a previous adapted criterion based on *P. falciparum* infection, as previously published by our group [6, 35]. Raw data used in all the analyses are concatenates in S1 File.

### Laboratory Measurements

Plasma levels of cytokines interleukin (IL)1-β, IL-4, IL-6, IL-10, IL-12p70, interferon (IFN)-γ, tumor necrosis factor (TNF)-α, and of C-C motif chemokine ligand (CCL)2, CCL5, C-X-C motif chemokine ligand (CXCL)9, and CXCL10 were measured using the Cytometric Bead Array -CBA^®^ (BD Biosciences Pharmingen, San Diego, CA, USA), according to the manufacturer’s protocol. The measurements of aspartate amino- transferase (AST), alanine amino-transferase (ALT), total bilirubin, direct bilirubin, creatinine, fibrinogen and C-reactive protein (CRP) were performed at the Pharmacy School of the Federal University of Bahia and at the clinical laboratory of *Faculdade São Lucas*, Brazil.

### Adaptation of Molecular Degree of Perturbation

The molecular inflammatory perturbation is based on the molecular degree of perturbation (MDP) method, used and recently published by our group [24–28]. In the present study, we inputted the plasma concentrations of a defined set of biomarkers based on previously published studies from our group in malaria pathogenesis [15,29,35,50–54]. Thus, herein, the average plasma concentration levels and the standard deviation of a baseline reference group (endemic controls) were calculated for each biomarker. The MDP score of an individual biomarker was defined by taking the differences in concentration levels from the average of the biomarker in the reference group divided by the reference standard deviation. Essentially, the MDP score represents the difference, by number of standard deviations, from the healthy control group. The equation used to calculate MDP in the present study is shown below:

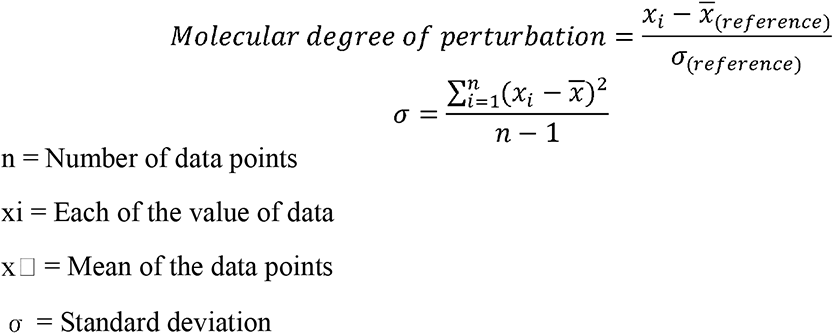

In this study we applied the MDP scoring system using data on 21 biomarkers measured from three distinct groups of patients, with symptomatic vivax infection, asymptomatic vivax infection and uninfected endemic controls. The MDP was filtered by the absolute MDP scores below 2 modules and by the sum of all accumulated MDP deviations. To identify samples implicated in ‘perturbation’, all values above the cutoff of the average MDP score plus 2 standard deviations of the reference group were considered “perturbed”. In additional analyses, the median value of ssMDP in symptomatic patients was calculated (value=48.11), and individuals exhibiting values above this threshold (top 50% symptomatic participants) were considered highly perturbed.

### Statistical Analysis

Descriptive statistics were performed to characterize the study population. Continuous variables were tested for Gaussian distribution using the D’Agostino-Pearson test. No variables exhibited normal distribution. The median values with interquartile ranges were used as measures of central tendency and dispersion, respectively. The Kruskal– Wallis test with the Dunn’s multiple-comparison were used to compare continuous variables whereas the Pearson’s chi-square test was used to compare variables displayed as percentages. Hierarchical cluster analyses (Ward’s method) of log10 transformed and z-score normalized data were employed to depict the overall expression profile of indicated biomarkers in the study subgroups. All comparisons were pre-specified and two-tailed. Differences with p-values below 0.05 after Holm-Bonferroni’s adjustment for multiple comparisons were considered statistically significant. Profiles of correlation between biochemical parameters were examined using network analysis of the Spearman correlation matrices. Correlations with p-value<0.05 were included in the network visualization. Spearman rank values (rho) were used to describe the strength of correlations between viral load from participants with current chronic infection and the MDP score of inflammatory parameters of these participants.

A Kaplan-Meier curve adaptation [33] was employed to test the probability of being highly perturbed according to the number of previous malaria episodes and age. A discriminant model using sparse canonical correlation analysis (CCA) was employed to assess whether inflammatory biomarkers, tissue damage biomarkers and inflammatory and tissue damage biomarkers could characterize the symptomatic participants. One polynomic (multinomial) logistic regression model was carried out using the variables with P-value <0.02 in the Mann-Whitney test between symptomatic and asymptomatic vivax participants, as showed in Table S1, to detect variables, epidemiologic or laboratorial, associated with symptomatic diseases.

Based on the mathematic formulas employed to visualize space-time deformity based on the general relativity theory proposed by Albert Einstein in 1915, we adapted the application of the proposed model to assess the overall impact of variables used here on homeostasis (defined as space-time membrane). In this model, homeostasis was defined by a plain p-brane which may be subjected to different degrees of deformities depending on the perturbation (distance from normal, which is calculated through the MDP scores) caused by each inflammatory marker, tissue damage markers and from the pathogen itself (parasitemia). The calculations and plots were build using the package *Shapes* from R.

The statistical analyses were performed using GraphPad Prism 8.0 (GraphPad Software, La Jolla, CA, USA), JMP 14.0 (SAS, Cary, NC, USA) and R statistical software.

## Results

### Characteristics of Study Population

Overall, study participants with symptomatic malaria were significantly younger when compared with those with asymptomatic *P. vivax* infection or uninfected controls (median age 29 [IQR 19-42]; 44 [IQR 34-50]; 39 [IQR 25-50] years, respectively, P<0.001). No differences were noted in regard to sex distribution (P=0.43). Of note, individuals with asymptomatic malaria referred a higher number of previous malaria episodes (100%, P<0.001). Also, statistical discrepancies were found in the time living in the endemic area, with, on average, fewer years reported by symptomatic patients (P<0.001). Noteworthy, the vast majority of MDP values calculated with the biomarkers measured in the study exhibited significant differences among groups. Additional findings are detailed in the S1 Table. Using a multivariate analysis, based on a stepwise model (as described in Methods), we confirmed that aging, presenting with more than ten years living in endemic area and increased number of previous malaria episodes were protective factors against occurrence of symptoms during the current malaria episode in the adjusted model, whereas the increases in the global inflammatory disturbance, measured by ssMDP, was a factor independently associated with the symptomatic malaria (S1 Figure panel A).

### Patients with symptomatic vivax malaria exhibit a higher degree of systemic inflammatory perturbation

To assess potential differences in the inflammatory activation according to the presence or absence of symptoms in vivax infection, we employed the molecular degree of perturbation (MDP, as described in Methods) in each study subpopulation (Fig 1). Individuals presenting with symptomatic vivax malaria exhibited higher ssMDP score values than those with asymptomatic infection or the uninfected control groups (Figs 1A and 1B). In addition, ssMDP values were also higher in asymptomatic malaria individuals than in those from the endemic control group (Fig 1B). These results reinforce the idea that the presence of *P. vivax* infection is associated with overall disturbances in the systemic inflammatory profile which degree is proportional to the clinical presentation.

**Figure 1:**
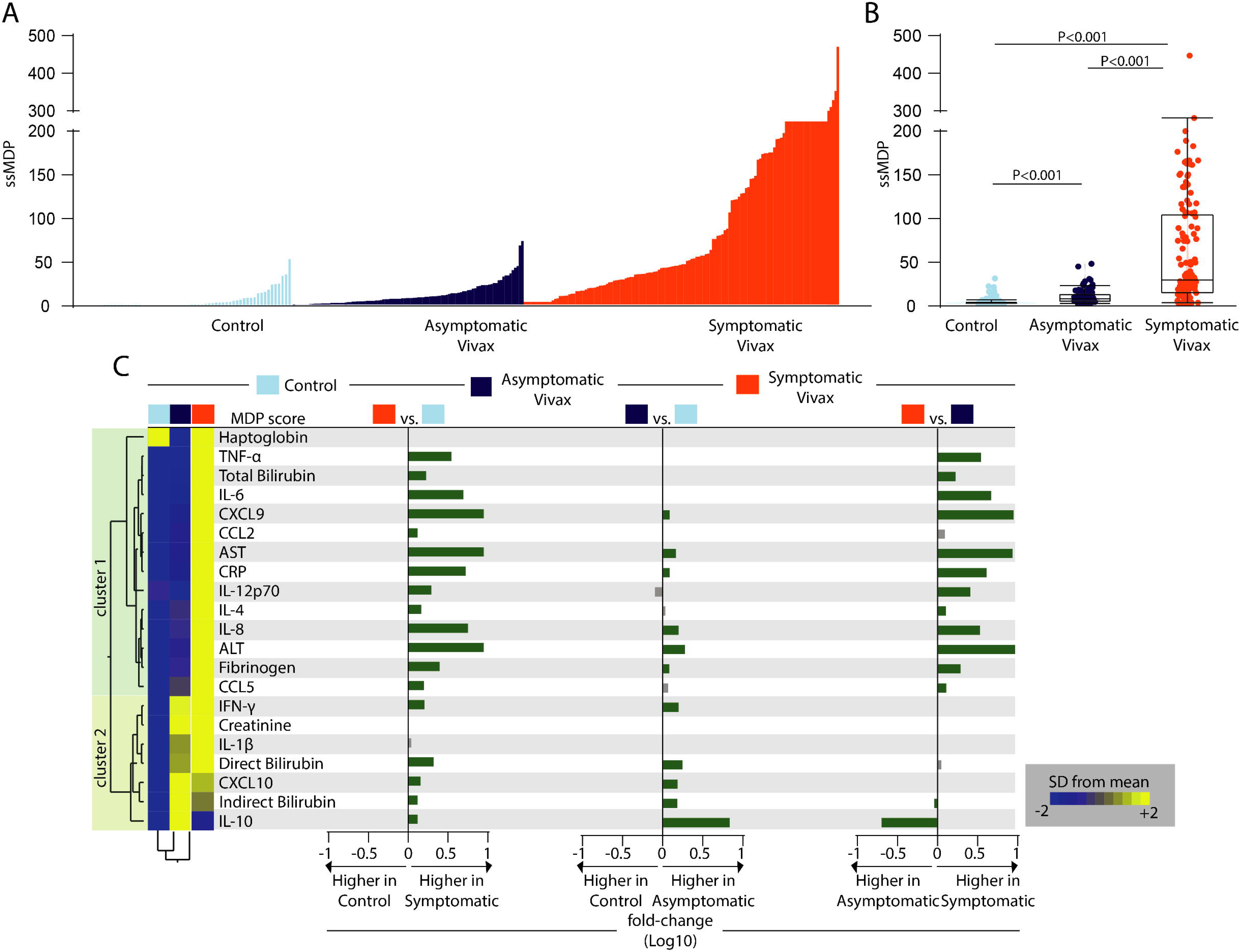
Higher degree of inflammatory perturbation associated with symptomatic vivax malaria. (A) Histograms show the molecular degree of perturbation (MDP) score values relative to each study group, as indicated. (B) Box plots represent the MDP score distribution between study groups, with median and interquartile range values. Data were compared between the clinical groups using the Kruskal–Wallis test, with Dunn’s multiple comparisons ad hoc test. (C) Left panel: Median MDP values for each indicated parameter were Log10 transformed and z-score normalized. A two-way hierarchical cluster analysis (Ward’s method) was employed to depict overall trends in changes of the MDP values among the clinical groups. Dendrograms represent the Euclidean distance. Right panel: Average fold-difference values of the MDP scores for each marker were calculated between clinical groups. Green bars represent statistically significant differences estimated by the Mann–Whitney U test. P-values were adjusted for multiple comparisons using the Holm-Bonferroni method. Abbreviations (alphabetic order): ALT: alanine aminotransferase; AST: aspartate aminotransferase; CCL: C-C motif chemokine ligand; CXCL: C-X-C motif chemokine ligand; CRP: C-reactive protein; IFN: interferon; IL: interleukin; TNF: tumor necrosis factor.

We next evaluated whether the degree of inflammatory perturbation calculated for each marker could provide more granularity about the disease endotypes’ details. The mean MDP score values of each marker of inflammation were analyzed by an unsupervised hierarchical cluster. Using this approach, we identified two main clusters of markers (Fig 1C). The markers forming the first cluster tended to increase the MDP values in the group of symptomatic malaria, with remarkable high values of TNF-α, IL-6, CXCL9, AST, CRP, IL-8 and ALT. The second cluster of markers was composed by IFN-γ, creatinine, IL-1β, direct and indirect bilirubin, CXCL10 and IL-10, with heightened MDP values being observed in both the group of individuals with either asymptomatic or symptomatic malaria. Patients with symptoms had the highest levels, except for IL-10, which was higher in the asymptomatic malaria group. An additional analysis using a binary multivariate logistic regression model revealed that increased perturbation scores of IL-6, IL-8 and CRP were independently associated with occurrence of symptomatic malaria, whereas higher degree of perturbation of IL-10 and fibrinogen were protective factors against symptoms in *P. vivax* infection (S1 Figure panel B). The visual differences detected in the heatmap of the MDP values were confirmed using fold-difference calculations between the study groups (Fig. 1C). Likewise, when comparing symptomatic with the uninfected control group, only perturbations of haptoglobin, creatinine and IL-1β were not significantly increased. The magnitude of differences in the MDP of the individual markers was found to be lower between the groups of individuals with asymptomatic malaria or uninfected controls. Alternatively, although the majority of markers with significant perturbances were noted in symptomatic patients, the comparison of malaria groups also identified augmented expression of MDP levels of IL-10 and indirect bilirubin in asymptomatic individuals. These findings confirm that susceptibility to *P. vivax* malaria’s susceptibility may be reflected by disturbances in the concentrations of several markers involved in controlling systemic inflammation.

### Network analysis of inflammatory balance in *Plasmodium vivax* infection

To more precisely characterize the nuances and dynamicity of the systemic inflammatory perturbation detected in the presence of *P. vivax* malaria, and which was more pronounced in those with symptomatic infection, we employed a network analysis of MDP values (Fig 2), as previously published by our group in a series of other pathological conditions [24–28].

**Figure 2:**
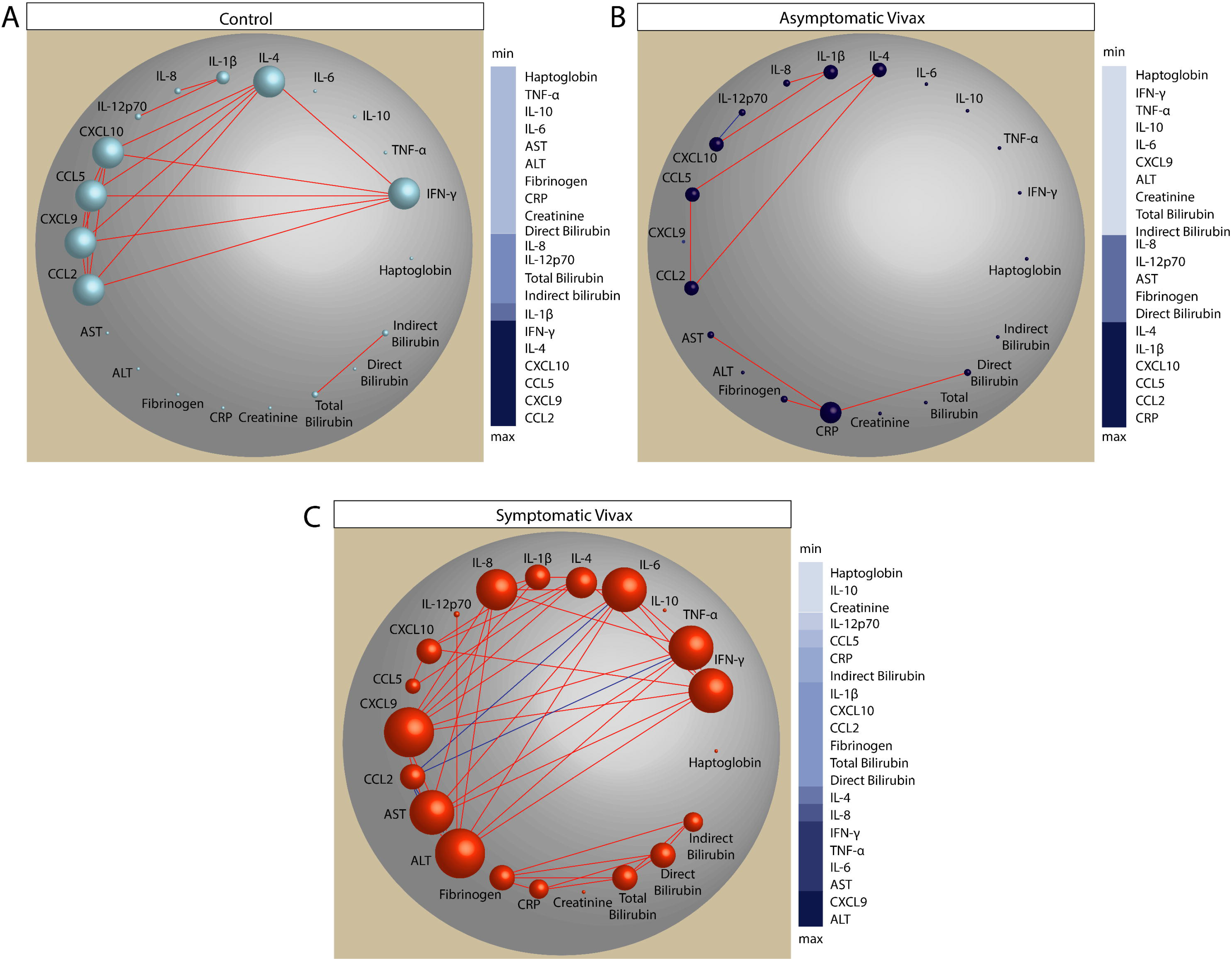
Network analysis of the MDP score values in the study groups. Spearman correlation matrices of the MDP score values in each study group - (A) controls; (B), asymptomatic vivax malaria; (C) symptomatic vivax malaria - were built and Circos plots were used to illustrate the correlation networks. Only significant correlations (p < 0.05 after adjustment for multiple comparisons) are shown. Each circle represents a different plasma parameter, and the size of each circle is proportional to the number of significant correlations. Lines represent the Spearman rank (rho) values. Red color infers positive correlation, whereas blue color denotes negative correlation. Node analysis heatmap shows the number of statistically significant correlations involving each marker per clinical group. Abbreviations (alphabetic order): ALT: alanine aminotransferase; AST: aspartate aminotransferase; CCL: C-C motif chemokine ligand; CXCL: C-X-C motif chemokine ligand; CRP: C-reactive protein; IFN: interferon; IL: interleukin; TNF: tumor necrosis factor.

Using MDP scores for each biomarker, as well as the overall MDP score (ssMDP), we found an important discrepancy in the network densities among the study groups. The group of symptomatic vivax participants exhibited the highest network density whereas asymptomatic infection displayed the lowest (Fig 2). Moreover, regardless of the clinical groups, most correlations were positive, meaning that increases in inflammatory perturbation of a given marker were frequently followed by increases in the disturbance levels of other inflammatory molecules (Fig 2). Curiously, the interferon pathway dominated the interactions (e.g., statistically significant correlations) in the control group, with CCL2, CXCL9, CCL5, CXCL10, IFN-γ and IL-4 showing up as the top highly connected markers (Fig 2A). In the group of asymptomatic malaria participants, the Spearman correlation matrices revealed a unique negative correlation, between the MDP values of CXCL10 and IL-12p70. The most highly connected markers were CRP, CCL2, CCL5, CXCL10, IL-1β and IL-4 (Fig 2B). Importantly, in the group of symptomatic vivax, we found two clusters of correlations: the first composed mainly by interactions between the inflammatory markers and ALT and AST; and the second composed by tissue damage markers (Fig 2C). Next, to verify the individual contribution to the global inflammatory perturbation, we employed a Spearman correlation between the ssMDP and the MDP level of each marker and found that the symptomatic group presented a higher number of significative markers associated with the global perturbation (S2 Figure). These observations argue that the appearance of symptoms in vivax malaria is associated with an intense disruption of the homeostasis, hallmarked by an increased overall degree of molecular perturbation as well as by a regulated process hallmarked by an intricate network of correlations of the individual markers.

### Time living in the endemic area influences the molecular degree of perturbation of individuals infected with *Plasmodium vivax*

We have previously demonstrated that persons living in malaria endemic areas from the Brazilian Amazon for more than 10 years tend to exhibit increased odds of developing asymptomatic malaria once infected with *P. vivax* [29]. Such findings agree with other investigations on *P. falciparum* infection in other regions of the globe [30, 31]. Here, to evaluate the impact of the time of residency in an endemic area on the molecular degree of inflammatory perturbation, we stratified the study participants in two groups: (i) one including those who lived less than ten years and (ii) a second group with persons who referred living more than ten years (Fig 3).

**Figure 3:**
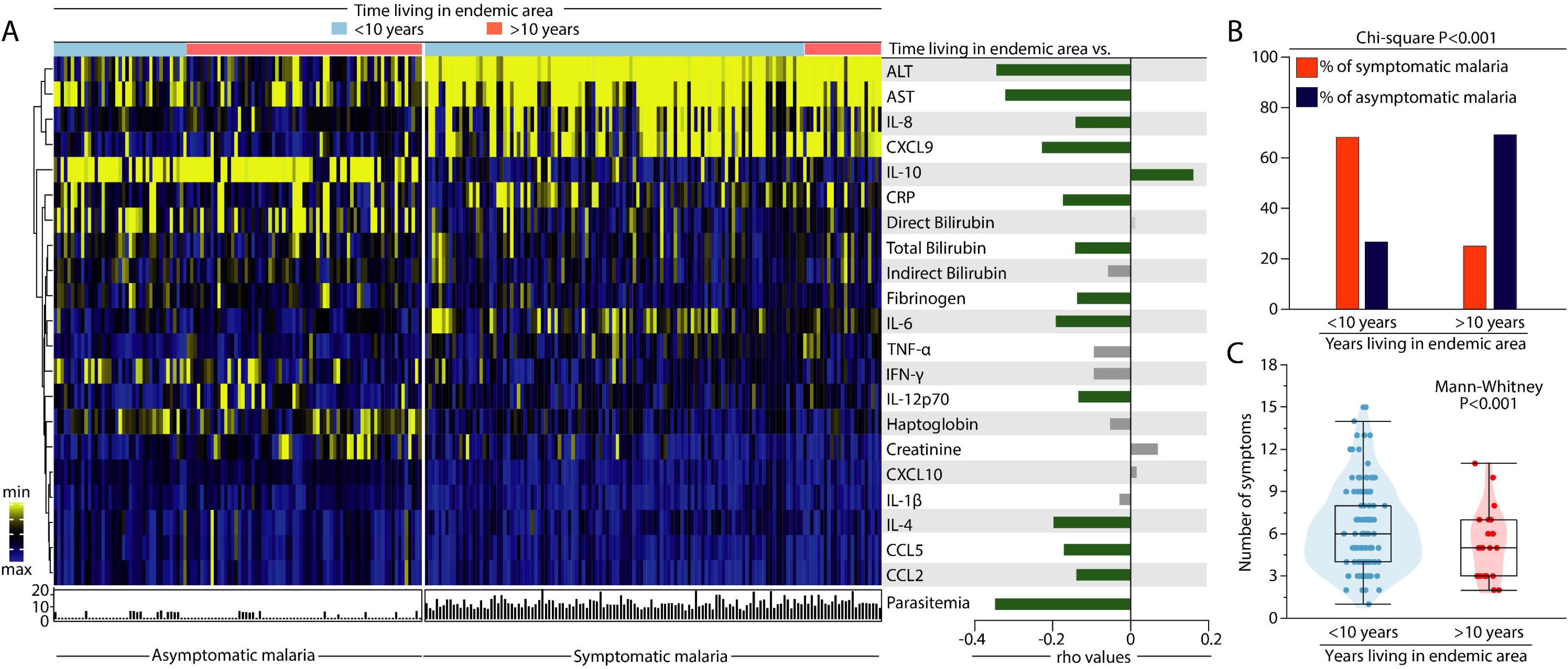
Patients with *Plasmodium vivax* malaria living longer in the endemic area display an attenuated systemic inflammatory disturbance. (A) Left panel: to build the heatmap, data were log-transformed and z-score normalized. A one-way hierarchical cluster analysis (Ward’s method) was employed to show the profiles of the molecular degree of perturbation of participants according to time living in an endemic area (patients were ordered according to time living in an endemic area). Patients were divided in two groups based on time living in the area (those who have lived for less than 10 years or for more than 10 years) and whether they presented with symptomatic or asymptomatic malaria. Right panel: Spearman rank test was used to assess correlations between the individual MDP score values of each parameter or parasitemia and time living in an endemic area (in years). Bars indicate the Spearman coefficient (rho) values. Green bars to the right indicate positive whereas to the left denote negative correlations with adjusted P-values <0.05. Grey bars indicate non-significant correlations. (B) The Pearson’s chi-square test was used to compare the frequency (percentage) of symptomatic and asymptomatic malaria participants according to the years living in an endemic area. (C) A violin plot was used to show the number of symptoms in the participants stratified according to years living in an endemic area. Data were compared using the Mann-Whitney U test. Abbreviations (alphabetic order): ALT: alanine aminotransferase; AST: aspartate aminotransferase; CCL: C-C motif chemokine ligand; CXCL: C-X-C motif chemokine ligand; CRP: C-reactive protein; IFN: interferon; IL: interleukin; TNF: tumor necrosis factor.

A hierarchical cluster analysis built upon a heatmap of z-score normalized MDP data on each individual marker and *P. vivax* parasitemia values failed to show a profile that could distinguish the individuals based on time living in the endemic area in both groups of malaria patients (symptomatic and asymptomatic malaria) (Fig 3A, left panel). A second investigative approach used Spearman correlation analysis between the MDP or parasitemia and time living in the endemic area in the entire subpopulation of *P. vivax*-infected individuals. Interestingly, the MDP scores from most of the markers as well as the parasitemia values were all negatively correlated with time living in the endemic area. One exception was IL-10 MDP values, which were positively correlated with time living in the area (Fig 3A, right panel). Of note, the perturbation of the liver damage parameters ALT, AST, total bilirubin, and also MDP values of IL-4, IL-6, IL-8, IL-12p70, CXCL9, CCL5, CCL2, CRP, fibrinogen and parasitemia levels showed an inverse correlation with the time living in the endemic area. Moreover, an expressive greater number of individuals with asymptomatic malaria was found to live more than 10 years in the area (Fig. 3B). Tolerance to malaria is tightly related to number of previous *Plasmodium* infections [20, 32] and people living for an extended time in endemic regions may be a higher odds of experiencing a higher number of *P. vivax* infections. We then tested whether time living in the endemic area would reflect the number of *P. vivax* symptoms. As expected, individuals living for longer periods in the endemic areas showed a lower number of symptoms in the current malaria episode. (Fig 3C).

### Impact of repetitive malaria episodes in the past on the degree of inflammatory perturbation in the current *Plasmodium vivax* infection

We next directly tested the hypothesis that individuals highly exposed to *P. vivax* and who experienced several previous malaria episodes would gradually learn how to modulate the immune activation and control the inflammatory responses that result in immunopathology, reducing the odds of developing more severe clinical presentations during the current infection. Initially, using Spearman correlation analysis, we found a general tendency to detect a negative correlation between the number of previous vivax malaria episodes and the MDP values of nearly all the markers evaluated (Fig 4A).

**Figure 4:**
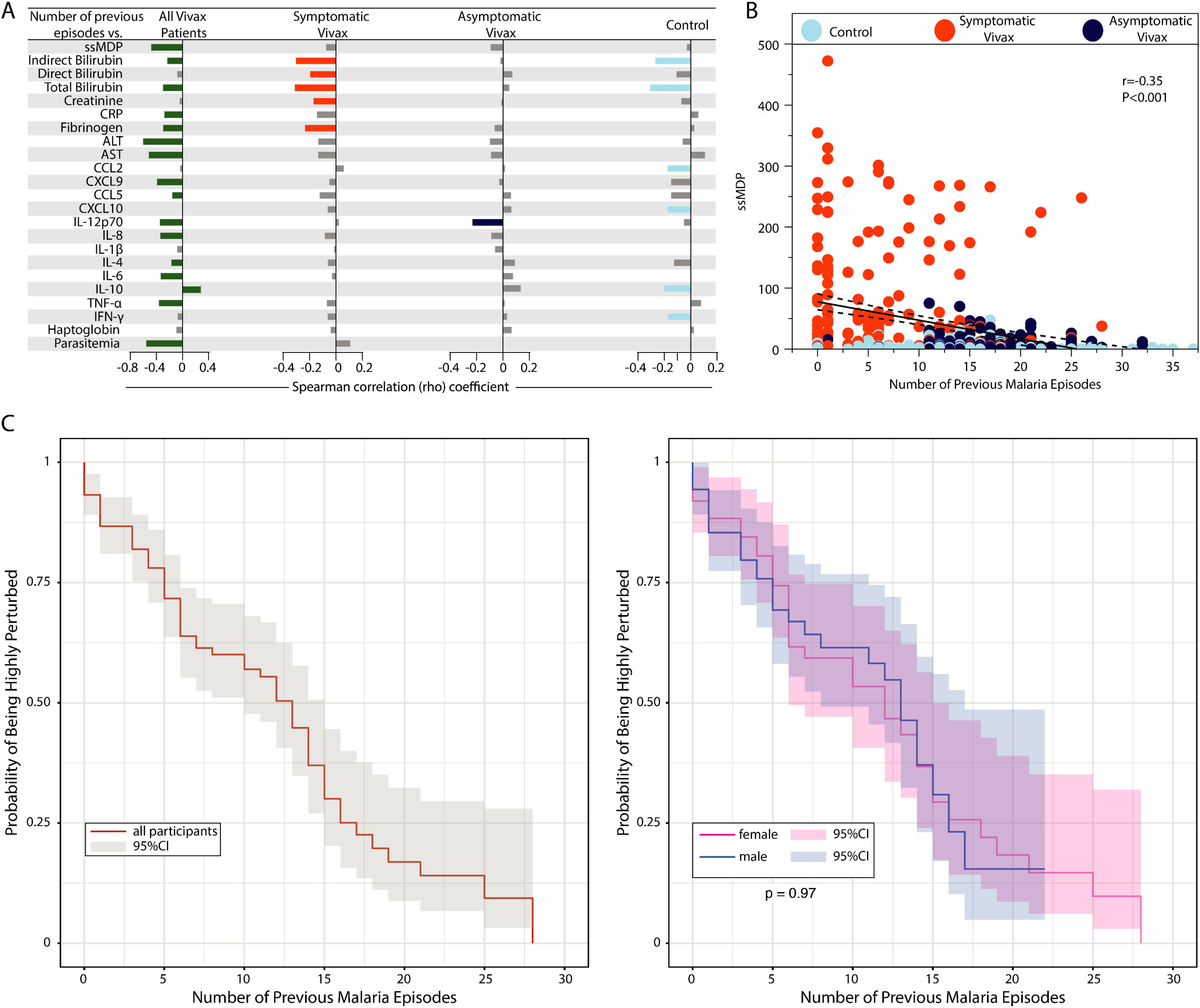
Heightened number of previous malaria episodes mitigates the inflammatory imbalance in the current *Plasmodium vivax* infection. (A) Spearman correlation between the number of previous malaria episodes and the global degree of inflammatory perturbation (ssMDP) of each parameter’s individual MDP values. Bars represent the Spearman rank coefficient value (rho). Colored bars (each clinical group is represented by a different color) infer correlations that presented adjusted P-value <0.05. (B) A scatter plot was employed to show the correlation between the number of previous malaria episodes and the ssMDP score values. (C) Probability of being highly perturbed (>2 standard deviations from control group) according to the number of previous malaria episodes in all participants (left panel) and participants grouped based on biological sex. The curves were compared using the Steger Method. Abbreviations (alphabetic order): ALT: alanine aminotransferase; AST: aspartate aminotransferase; CCL: C-C motif chemokine ligand; CXCL: C-X-C motif chemokine ligand; CRP: C-reactive protein; IFN: interferon; IL: interleukin; TNF: tumor necrosis factor.

Applying this analysis to all participants with malaria, we observed that only MDP levels of IL-10 were directly associated with the number of previous malaria episodes (Fig 4A). An interesting finding was the negative correlation between the number of previous episodes and parasitemia in all the participants with vivax malaria, independent of the presentation of symptoms (Fig 4A). In participants with symptomatic malaria, only negative correlations were detected, highlighting the influence of previous infection episodes on reducing perturbance of tissue damage markers, such as direct and indirect bilirubin, creatinine, and fibrinogen. This observation reinforces the idea that having a history of several previous *Plasmodium vivax* infections is related to a relative control of tissue damage (Fig 4A). Extending the approach to asymptomatic participants, we detected a unique inverse correlation between the number of previous episodes and IL-12p70 MDP values. Additionally, we found that even in the uninfected control group, history of previous malaria episodes was associated with reduced perturbation of indirect bilirubin, total bilirubin, CCL2, CXCL10, IL-10 and IFN-γ (Fig 4A). Importantly, when all malaria participants were analyzed, the global perturbation measured by ssMDP levels was lower proportionally relative to the number of previous malaria episodes (Figs 4A and 4B). Finally, a Kaplan-Meier survival curve demonstrated that increases in the number of previous malaria episodes resulted in a gradual lower probability of a study participant being considered highly perturbed (presenting with ≥±2SD of the overall ssMDP score value, as described in Methods), independent of biological sex (Fig 4C). Noteworthy, our analysis revealed that previous exposure to *Plasmodium* infections indeed seems to attenuate the immune activation of the current episode, reflecting the lower odds of experienced an unfettered, unbalanced, inflammatory response.

### Aging impacts the molecular degree of perturbation in *Plasmodium vivax* infection

We have previously suggested that there is an effect of aging in the modulation of systemic inflammation in *P. vivax* infection [7]. Here, we tested whether such an effect may be explained by the impact of aging on the molecular degree of inflammatory perturbation. Hence, Spearman correlation analysis revealed that, in general, increasing in age was associated with decreases in the MDP score values of the markers investigated in the study population (Fig 5).

**Figure 5:**
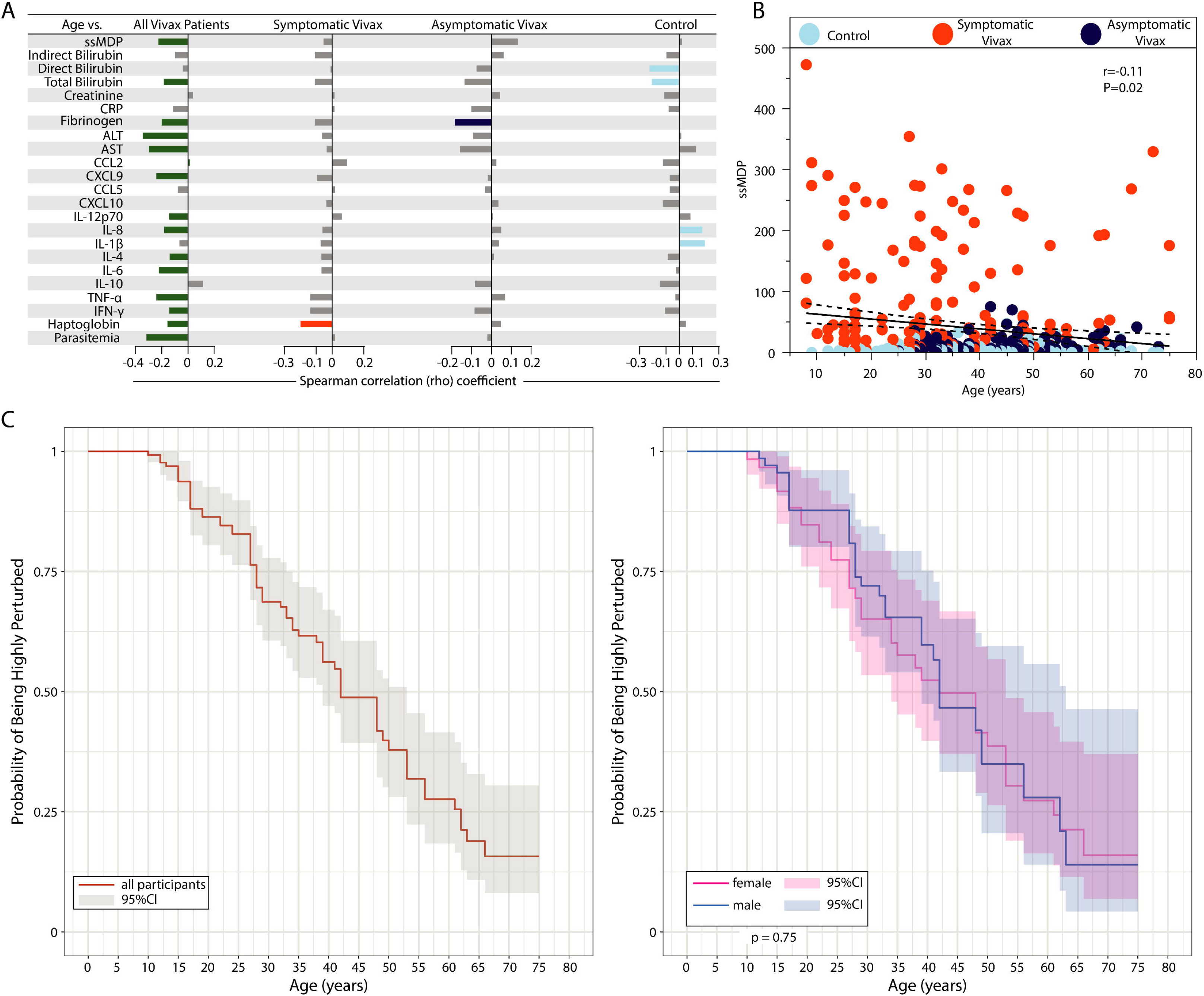
Aging is associated with diminished systemic inflammatory disturbance in vivax malaria. (A) Spearman correlation between age and the global degree of inflammatory perturbation (ssMDP) of the individual MDP values for each parameter. Bars represent the Spearman rank coefficient value (rho). Colored bars (each clinical group is represented by a different color) infer correlations that presented adjusted P-value <0.05. (B) A scatter plot was employed to show the correlation between age and the ssMDP score values. (C) Probability of being highly perturbed (>2 standard deviations from the control group) according to age in all participants (left panel) and in participants grouped based on biological sex. Abbreviations (alphabetic order): ALT: alanine aminotransferase; AST: aspartate aminotransferase; CCL: C-C motif chemokine ligand; CXCL: C-X-C motif chemokine ligand; CRP: C-reactive protein; IFN: interferon; IL: interleukin; TNF: tumor necrosis factor.

Considering only vivax participants, all the statistically significant correlations between aging and MDP score values were negative, meaning that increases in age were directly related to a reduction in the global inflammatory disturbance (ssMDP), in the individual degree of perturbation of fibrinogen, ALT, AST, TNF-α, IFN-γ, haptoglobin and also parasitemia (Fig 5A). Importantly, in the group of symptomatic malaria, aging was significantly related to decreased haptoglobin MDP values only, whereas in the group of asymptomatic malaria, aging specifically reduced the fibrinogen MDP values but not of other markers (Fig 5A). In the endemic control group, we found that aging was associated with increases in MDP scores of IL-8 and IL-1β and decreased MDP values of total and direct bilirubin. We next evaluated the impact of aging in the global inflammatory perturbation in all of our study participants and found a negative correlation (r=0.011; P=0.032) (Fig 5B). Further, increases in age were shown to proportionally reduce the probability of an individual being highly perturbed regardless of the biological sex (Fig 5C), suggesting that aging mitigates the inflammatory imbalance in *P.vivax*-infected individuals. Finally, a hierarchical cluster analysis performed based on the age and number of malaria episodes could not distinguish groups when all participants were inputted. However, it reinforced our results exhibiting the influence of aging and previous infections over more controlled inflammatory responses (reduced MDP values) and lower frequency of symptomatic malaria cases (S3 Figure).

### Effects of the inflammatory disturbance on the number of symptoms during *Plasmodium vivax* infection

To dissect the clinical effects of the inflammatory imbalance during *P. vivax* infection, we used a color map analysis, ordinating the participants based on the number of symptoms, to describe the symptomatology distribution in our study population (Fig 6A).

**Figure 6:**
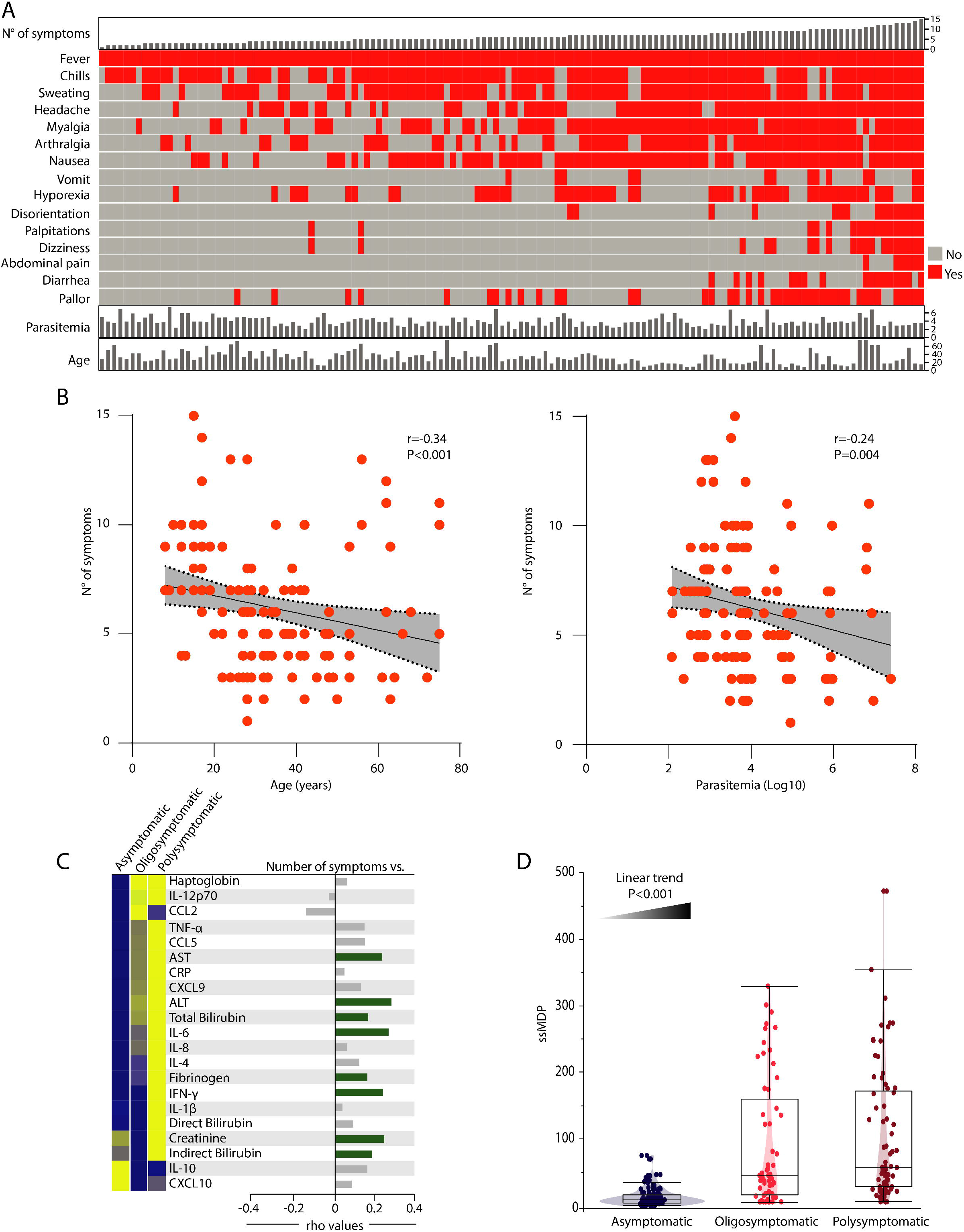
Impact of systemic inflammatory disturbance on the number of symptoms in *Plasmodium vivax* malaria. (A) Symptomatic *Plasmodium vivax* malaria patients were ordered according to the number of symptoms. A color map highlighting the detection of each symptom is shown. Parasitemia (log10) and age are shown as continuous variables on the bottom for each patient. (B) Spearman correlation between the number of symptoms and age (left panel) and Log10 parasitemia (right panel). Linear curves with 95% confidence intervals were used to illustrate trends in data variation. (C) Patients were stratified into three groups based on the presence of symptoms: Asymptomatic, Oligosymptomatic (≤5 symptoms) or Polysymptomatic (>5 symptoms). Left panel: Median MDP values for each indicated parameter were Log10 transformed and z-score normalized. A one-way hierarchical cluster analysis (Ward’s method) was employed to depict overall trends in changes of the MDP values among the clinical groups. Dendrograms represent the Euclidean distance. Right panel: Spearman correlation between the number of symptoms and the MDP score values for each indicated parameter. Green bars indicate statistically significant correlations (all positive after adjusted P-value <0.05). Grey bars denote a non-significant correlation. (D) Violin plot shows the distribution of the global degree of perturbation (ssMDP) in each subgroup of Plasmodium vivax patients. Data were compared using the Kruskal-Wallis test with a non-parametric linear trend posttest.

As expected, fever was the main symptom documented in *P. vivax*-infected participants, which was identified in all patients, followed by chills, sweating, headache, myalgia, arthralgia and nausea. We next employed Spearman correlation analyses and found a curious negative correlation between the number of symptoms and both age and parasitemia (r =-0.34, P<0.001; r=-0.24, P=0.004, respectively) (Fig 6B). Next, we divided our participants according to the number of symptoms, in asymptomatic (no symptoms), oligosymptomatic (less than six symptoms) and polysymptomatic (those who presented six or more symptoms) and employed a hierarchical cluster and a Spearman correlation analysis to investigate the potential effect of the inflammatory activation (measured through MDP score values of the markers) on symptomatology in *P. vivax* infection (Fig 6C). A clear distinction in the MDP profiles of the markers was observed in the unsupervised hierarchical cluster between asymptomatic, oligosymptomatic and polysymptomatic groups (Fig. 6C). Overall, the group of polysymptomatic individuals exhibited relatively heightened inflammatory perturbation values, distinguished from the other groups by expression of almost all markers when compared with asymptomatic participants. Precisely, increased IFN-γ, IL-1, creatinine, direct and indirect bilirubin MDP scores in comparison with the oligosymptomatic group (Fig 6C). Asymptomatic participants, on the other hand, showed a tendency to express higher MDP values of IL-10 and CXCL10, which were clearly lower than the oligo and polysymptomatic groups (Fig 6C). Furthermore, Spearman correlation analyses were performed to investigate the association between the number of symptoms and the degree of inflammatory perturbation of the markers indicated. This approach revealed that the number of symptoms was associated mainly with increases in the degree of perturbation of tissue damage markers, such as AST, ALT, total and indirect bilirubin, fibrinogen and creatinine (Fig 6C). Additionally, we found that the global molecular perturbation measured by ssMDP values exhibited a gradual tendency to increase in the participants according to the augmented number of symptoms (Fig 6D). Our findings argue that the degree of inflammatory disturbance in peripheral blood of *P. vivax* malaria patients is directly related to the clinical presentation severity.

### Perturbation in concentrations of tissue damage markers, rather than of immune activation molecules characterizes symptomatic *Plasmodium vivax* malaria

Among the biomarkers investigated in the present study, there were mostly two classes of molecules: (i) those that more closely infer inflammatory activity and (ii) those intrinsically related to tissue damage/injury (Fig 7A).

**Figure 7:**
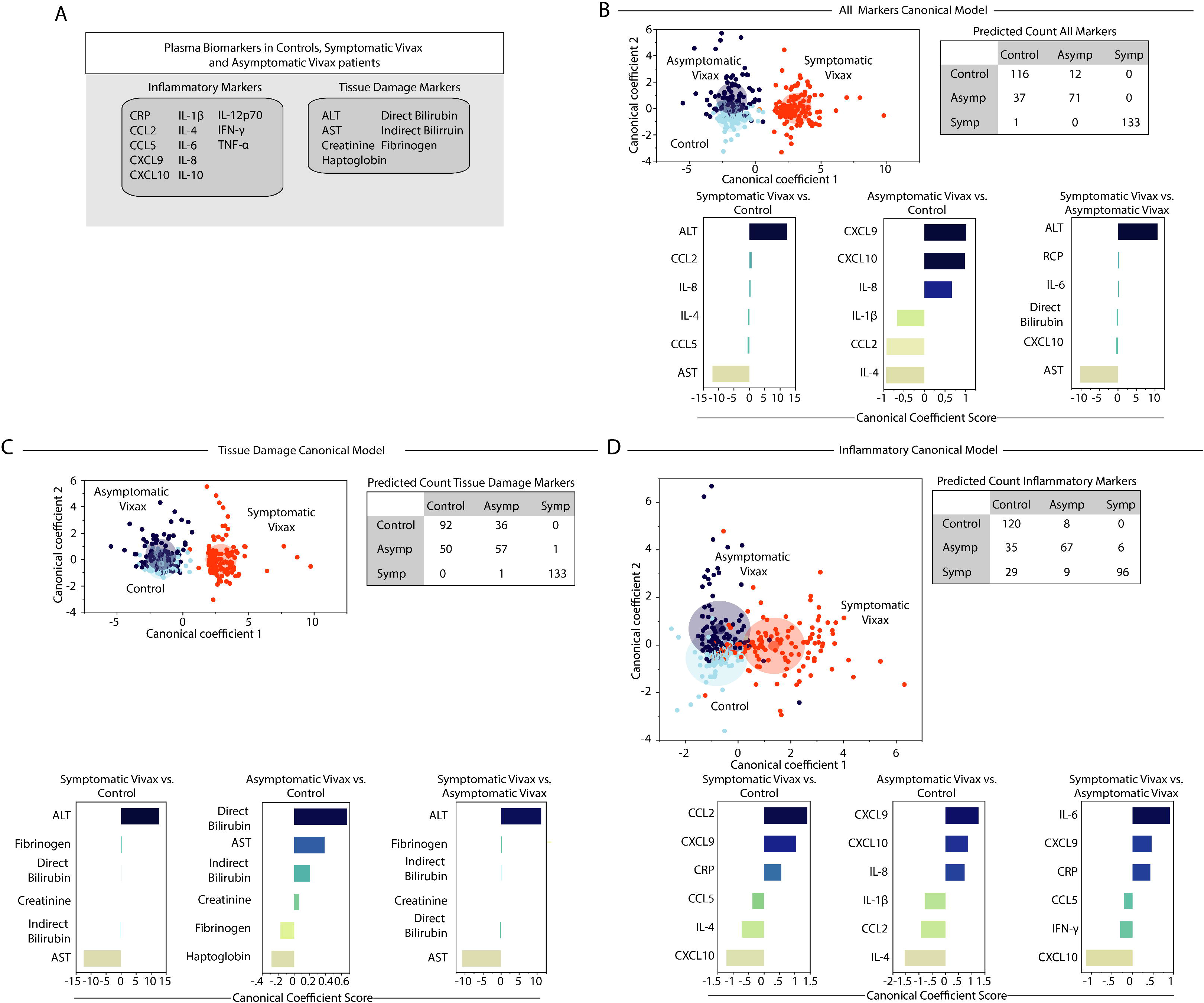
Using canonical correlation analysis of the degree of perturbation of inflammatory or tissue damage markers to characterize vivax malaria. (A) In an exploratory approach, a sparse canonical correlation analysis (sCCA) was used to test whether the indicated clinical groups could be distinguished based on: (B) the overall perturbation profile of all the markers measured, or combinations of (C) tissue damage markers or (D) inflammatory markers only. Lower panels show the canonical coefficient scores calculated to identify the biomarkers responsible for the distinctions between groups in each sCCA model as indicated. Abbreviations (alphabetic order): ALT: alanine aminotransferase; AST: aspartate aminotransferase; CCL: C-C motif chemokine ligand; CXCL: C-X-C motif chemokine ligand; CRP: C-reactive protein; IFN: interferon; IL: interleukin; TNF: tumor necrosis factor.

To more precisely define which class of markers is more relevant to explain the occurrence of symptoms in individuals infected with *P. vivax*, we used a discriminant algorithm based on sparse canonical correlation analysis (sCCA). We performed three models (i) one with all the markers (Fig 7B), (ii) a second model with only the tissue damage markers (Fig 7C) and (iii) a final model including only inflammation-related markers (Fig 7D). In all three models, discrimination of symptomatic participants from asymptomatic and endemic controls was detected with a high degree of accuracy (Fig 7B-D). As expected, differentiation between asymptomatic malaria and uninfected controls was difficult, with many misclassifications seen in the models. We next assessed the canonical coefficients of each model, which is a statistical strategy to rank the markers that most contributed to the discriminant model. This approach found that AST and ALT were the top markers associated with symptomatic participants (Fig 7B). In the second model, canonical coefficient values of tissue damage markers, ALT and AST also emerged as the top markers that could identify individuals with symptomatic malaria, whereas direct bilirubin and haptoglobin the top damage markers associated with the distinction between asymptomatic malaria and uninfected controls (Fig 7C). Finally, we used only the inflammatory markers (Fig 7D) and observed a relevant decrease in the discrimination power, with 38 misclassifications in the group of symptomatic malaria. Using information from the canonical coefficients, we found that the IFN pathway showed a relevant role in the discrimination of symptomatic malaria and controls, with CCL2 followed by CXCL10 and CXCL9 as the top markers. CXCL9 and IL-4 were the top markers when we analyzed asymptomatic malaria vs. controls, whereas IL-6 and CXCL10 were highlighted when symptomatic vs. asymptomatic malaria patients were compared (Fig 7D). In the context of *P. vivax* malaria, these observations reinforce the idea that alterations in circulating concentrations of tissue damage-associated markers more intrinsically relate with the appearance of symptoms than the imbalance of the immune activation alone.

### Characterizing disease tolerance in *Plasmodium vivax* malaria

We further tested whether disease tolerance could be observed and characterized in the study participants infected with *P. vivax*. To do so, we examined the systemic inflammation assessed through the ssMDP values in the context of *P. vivax* parasitemia and the number of symptoms. First, we employed a hierarchical cluster with z-score normalized values to depict the overall distribution of these three selected parameters in the group of patients with symptomatic malaria. Interestingly, the combination of the ssMDP, number of symptoms, and *P. vivax* parasitemia was not able to completely segregate patients with severe malaria from those with non-severe malaria (See definitions used to classify patients based on malaria severity in Methods) (Fig 8A).

**Figure 8:**
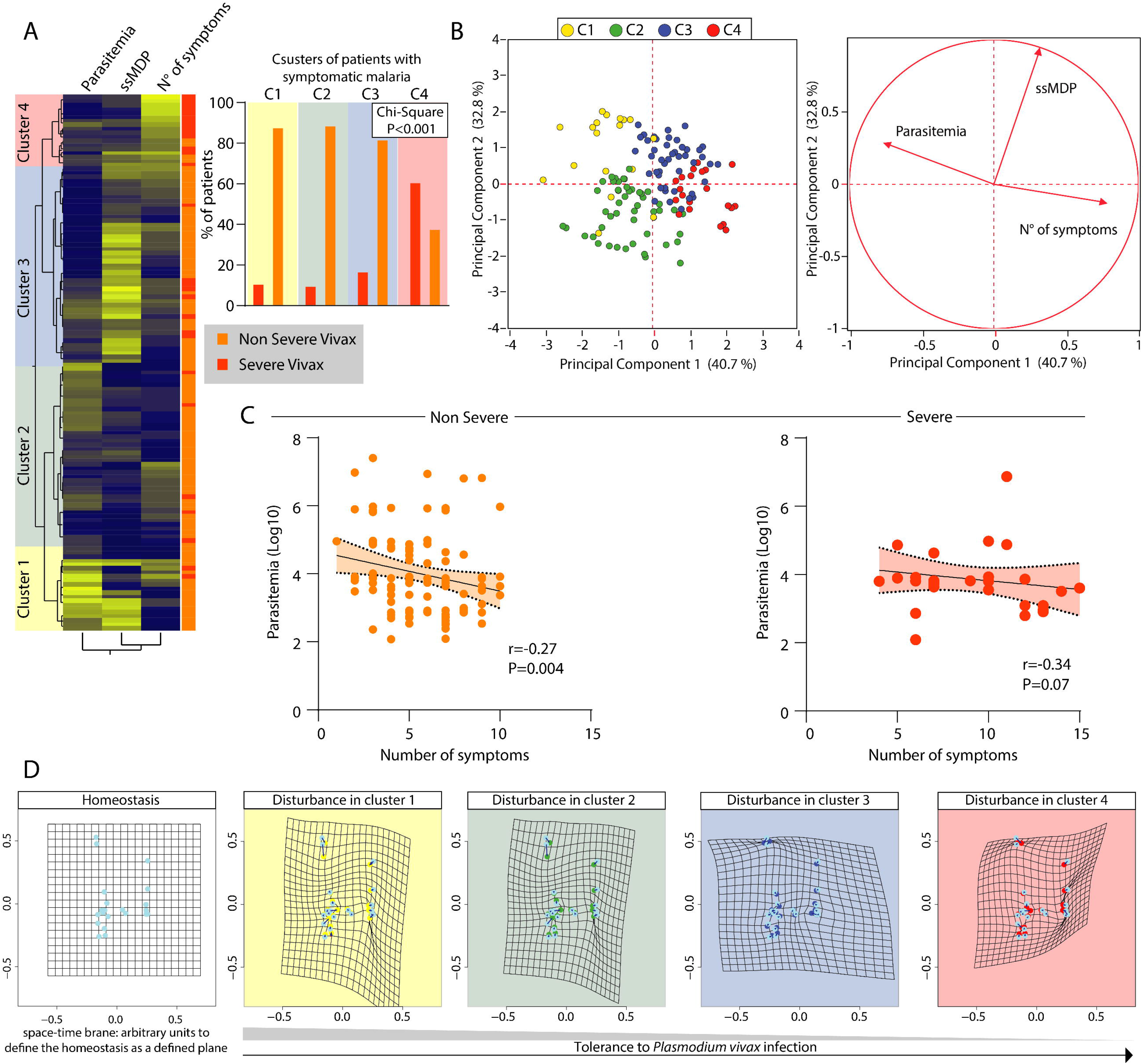
Characterizing disease tolerance in *Plasmodium vivax* malaria. (A) Left panel: A two-way hierarchical cluster analysis was employed to characterize the overall profiles of parasitemia, systemic inflammatory disturbance (ssMDP) and the number of symptoms in Plasmodium vivax patients presenting with either severe (n=28) or non-severe disease (n=106). Right panel: A Chi-Square test was used to test the proportion of severe vivax patients in each cluster identified. (B) A principal component (PCA) model (left panel) with a vector plot (right panel) was used to demonstrate the influence of each indicated parameter on the distribution of the data. Colors indicate the clusters identified in panel (A). (C) Patients were segregated according to malaria severity and a Spearman correlation analysis was performed to identify the association between parasitemia and number of symptoms (N° of symptoms). (D) A space-time deformity model based on the relativity theory of gravity was employed to visualize the impact of each plasma marker and tissue damage marker on disturbance of the homeostasis (defined as space-time brane in the control group). See Methods for details on the mathematical model. Each dot represents a marker, and groups of patients were separated based on the cluster analyses shown in (A).

Moreover, the clustering analysis revealed four main clusters of study participants. The first cluster was composed predominantly by patients with non-severe malaria. Such cluster was characterized by the highest parasitemia and variable ssMDP values, with a low number of symptoms (Fig 8A). The second cluster was also composed mainly by non-severe malaria participants and exhibited a relatively low value for all three parameters evaluated (Fig 8A). The third cluster displayed a slight increase in the frequency of participants with severe malaria and was marked by augmented levels of global molecular perturbation, despite lower parasitemia values and the number of symptoms (Fig 8A). Finally, the fourth cluster was constituted by a majority of participants with severe malaria. This latter cluster exhibited more commonly a higher number of symptoms despite the relative lower ssMDP values and parasitemia levels (Fig 8A). These results suggest that the fourth cluster of study participants included less tolerant to *P. vivax* malaria patients. A principal component analysis (PCA), using the same parameters inputted in the hierarchical clustering, validated the idea that the number of symptoms, systemic inflammation assessed by ssMDP, and parasitemia were all independently influencing the distribution of the participants with symptomatic malaria into different subgroups of individuals. The PCA groups were similar to the clusters identified in the clustering analysis (Fig. 8B). Importantly, correlation analyses revealed an inverse linear association between parasitemia and the number of symptoms in individuals with non-severe malaria but not in those with severe malaria (Fig 8C). We next employed statistics previously used to assess space-time deformity by gravity following the general relativity theory (as described in Methods) to visualize the impact of each inflammatory and tissue damage marker on homeostasis disturbance. The disturbance is defined here by comparison to the space-time brane calculated for the uninfected endemic control group. Using this approach, the degree of deformity in the space-time brane infers the degree of homeostatic disturbance. The analyses demonstrated that the fourth cluster of malaria, mainly composed of severe malaria patients, exhibited the highest homeostatic disturbance (Fig 8D). As shown in Fig 8D, there was a progressive disturbance of the homeostasis through clusters 1 to 4, revealing a remarkable tolerance pattern, in which patients from the first cluster exhibited more robust tolerance to the disease.

### Identifying the Determinants of Disease Tolerance in *P. vivax* Infection

Spearman correlation matrices between the MDP values of the inflammatory or tissue damage markers and the number of symptoms and parasitemia values were employed to dissect the factors that could directly or indirectly influence tolerance in vivax malaria patients (Fig 9).

**Figure 9:**
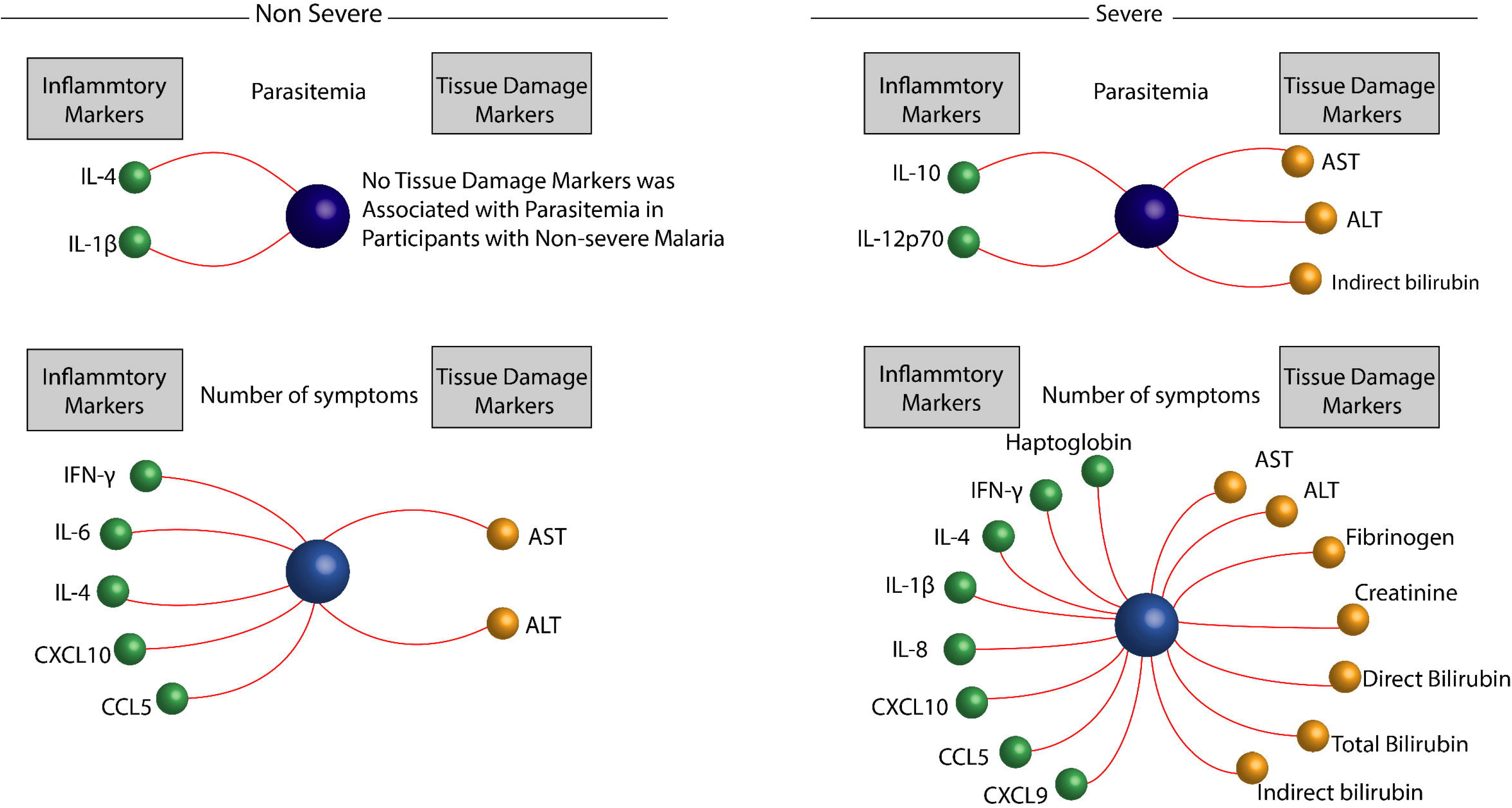
Identifying the factors associated with disease tolerance in *Plasmodium vivax* malaria. A Spearman correlation rank matrix analysis was used to identify the specific inflammatory or tissue damage markers which degree of disturbance is associated with parasitemia or number of symptoms (the latter are represented as central spheres in the network graphs) in patients non-severe or severe P. vivax as indicated. Each line represents a statistically significant correlation (p<0.05) after adjustment for multiple measurements. Among the statistically significant correlations detected, only positive associations were found.

In participants with non-severe vivax malaria, our analysis revealed that only MDP levels of IL-1β and IL-4 were positively correlated with parasitemia. MDP levels of IFN-γ, IL-4, IL-6, CXCL10, CCL5, and liver parameters ASL and ALT showed to be positively correlated with the number of symptoms (Fig 9). Extending our approach to participants with severe malaria, we found that MDP levels of IL-10, IL-12p70, AST, ALT, and indirect bilirubin were directly associated with parasitemia (Fig 9). Importantly, the number of symptoms in those presenting severe vivax infection was associated with increases of several MDP levels of both inflammatory and tissue damage markers.

## Discussion

Knowledge about disease tolerance in humans remains poorly elucidated, in part due to the difficulty of establishing a reliable measure of systemic inflammation. Here, to fill this gap, we used the molecular degree of perturbation (MDP) approach, a statistical tool that led us to infer the global inflammatory status, and which has been recently proposed by our group to help to understand the pathophysiology of a range of infectious and non-infectious diseases [24–27,33,34]. We evaluated the effects of several epidemiological characteristics in inflammatory responses to *P. vivax*, dissecting the disease tolerance to vivax malaria and identifying the molecular factors associated with the phenomenon.

Using the MDP, we found that regardless of the symptomatology, *P. vivax* infected participants exhibited a higher degree of inflammatory perturbation than the control groups. Importantly, symptomatic patients displayed the highest global MDP scores. In fact, we have previously reported that the clinical manifestation in *P. vivax* infection was strongly associated with an inflammatory imbalance in peripheral blood [35]. Here, we also identified a non-surprising pattern of inflammatory imbalance present in all the infected participants. Such a pattern comprises a higher degree of perturbation of CXCL9, CXCL10, IL-8, IL-10, IFN-γ, AST, ALT, CRP, Fibrinogen, direct and indirect bilirubin concentrations. Moreover, only infected symptomatic patients had TNF-α, total bilirubin, and IL-6 increased perturbation. These biomarkers have been extensive explored in the context of malaria pathophysiology. IFN-γ and CRP have been recently identified as an inflammatory signature associated with a higher risk of mortality in *P. vivax* infection[6]. Polymorphisms in TNF-α and IL-6 genes [36], as well higher levels of these markers and of CXCL10 [37] were linked to malaria clinical outcomes and pro-inflammatory activation in response to *P. vivax* [38]. IL-10 has a fundamental role in response to *Plasmodium* infections, directly affecting dampening the inflammation-driven cytokine storm, modulating the immune response, and limiting the tissue damage, reducing odds of severe malaria [39–41]. Using the MDP assessment in malaria patients led us to define the nuances of the inflammatory imbalance associated with *P.vivax* infection.

Conceptually, inflammation is a dynamic process aimed at reestablishing tissue homeostasis. In some infectious diseases, such as malaria, the balance between immune activation, regulatory cytokine responses and degree of tissue damage is a critical determinant of disease outcomes [42–44]. We explored the dynamicity of the *P.vivax* infection using the Spearman correlation matrices [24,25,39,45], and found a higher density of correlations among the participants with symptomatic malaria, with predominance of positive correlation among the degree of perturbation of the biomarkers. Despite that, in participants with symptomatic malaria, the degree of CCL2 perturbation exhibited two negative correlations, vs. IL-6 and TNF-α. Moreover, ALT was the top node in the matrix calculated for symptomatic malaria, highlighting the relevance of liver inflammatory reaction in *P.vivax* infection [46, 47]. Importantly, different from our previous publication using absolute concentration values of inflammatory markers, where we had identified a highest number of correlations in matrices from asymptomatic patients [39], here, by imputing MDP values instead, we found that this group of participants exhibit the lowest network density, suggesting that the absence of symptoms in response to *P. vivax* is associated with a potential uncoupling of the perturbed inflammatory response, probably due to lack of immune activation in peripheral blood. A curious finding was the relative dissociation between tissue inflammatory damage markers and immune factors. This dissociation argues that, except for the liver, the symptoms caused by *P.vivax* are strongly associated with the cytokine storm in response to infection and may be also linked to local tissue reaction, assessed by measuring levels of liver transaminases for example, but without direct interaction between these MDP levels. This dissociation can be a consequence of the immune evasion mechanisms presented by the parasite, which alter the red blood cell (RBC) structure through expression and export of molecules encoded by plasmodium genome [48]. The interaction between the changed RBC and the host immune cells hinders the plasmodium recognition and enables the tissue damage directly mediated by the pathogen.

Epidemiological aspects have largely been described to influence clinical presentation and outcomes in malaria [7,30,31,48]. Many these effects are also related to the profile of the immune response. In continuous exposure to *P.vivax*, reflected throughout time living in endemic area and age, the host develops clinical immunity against *Plasmodium.* Still, a controlled inflammatory activation can limit the parasite burden [7] without major perturbations in homeostasis, dampening symptomatology. Here, we showed that repetitive *P. vivax* infection in the past gradually mitigates the inflammatory activation in later infection episodes. The indications of this phenomenon are several. First, we found a robust negative correlation between the time living in an endemic area and the MDP values for all markers measured, except for IL-10 MDP levels. Our results also revealed a higher incidence of symptomatic individuals and fewer symptoms among those who reported living less than ten years in the endemic region. Next, we showed negative correlations between the number of previous malaria episodes and the inflammatory imbalance detected during the current infection, except for, again, IL-10 MDP levels. It is important to note that we found a tendency to decrease systemic inflammation and a lower probability of being highly perturbed in individuals with a greater number of previous malaria episodes.

Regarding age, we found that aging is associated with less intense inflammatory activation. Reinforcing the role of hyper-inflammatory status in symptomatology, we demonstrated that the number of symptoms was positively associated with increased MDP levels. Precisely, levels of tissue damage markers, such as AST, ALT, bilirubin, fibrinogen, and creatinine, and the pro-inflammatory markers IL-6 and IFN-γ Additionally, the ‘global’ systemic inflammation, measured by ssMDP levels, was higher in polysymptomatic participants. Our observations reported here suggest that older individuals who lived for long time in malaria-endemic area and had been exposed to several Plasmodium infections in the past tend to tolerate the current *P. vivax* infection through a decrease in perturbation of all pro-inflammatory and tissue damage markers, and increase in MDP levels of IL-10, which may limit tissue damage and minimize sickness [39–41].

Finally, our study tried to dissect the determinants of disease tolerance to *P. vivax* infection. Using data on parasitemia, ssMDP values, and the number of symptoms, we were able to evaluate the interplay between the main factors associated with unfavorable outcomes in an infectious disease: (i) pathogen load; (ii) host response to infection and (iii) clinical consequences of this interaction. Our analysis reveals that the main factor associated with severe cases in *P. vivax* infection was the number of symptoms, demonstrating a robust effect on homeostasis disruption. Of note, these participants exhibited relatively lower parasitemia and ssMDP values, arguing that some persons present severe vivax cases regardless of pathogen burden and inflammatory activation, being little tolerant to infection. In converse, we identified a subgroup of individuals who presented with higher parasitemia and augmented ssMDP values but lower symptomatology and number of severe malaria cases. This latter group of individuals may be activating the immune response to counteract the parasite load, but without resulting in substantial tissue damage and symptomatology. We also found another group of *P. vivax*-infected study participants who seemed to have controlled infection without inducing substantial inflammatory disturbance, and presented with low parasitemia, lower ssMDP values and few symptoms. Importantly, we found that persons with severe vivax malaria lost the effective control of parasitemia resulting in tissue damage and symptoms, exemplified here by absence of a statistically significant correlation between parasitemia and number of symptoms.

Our final results disclosed the determinants of diseases tolerance in *P. vivax* infection. To do that, we employed networks inputting correlations between degree of molecular perturbation of tissue damage or inflammatory markers with parasitemia and number of symptoms. We described that higher parasitemia in severe vivax malaria was associated with a more robust tissue response, and that the degree of symptomatology was directly related to degree of perturbation in cytokine concentrations. Such scenario may significantly contribute to tissue damage through systemic inflammatory activation and also tissue inflammation, reinforcing our previous results reported above. Nevertheless, in non-severe cases, parasitemia was not associated with MDP of tissue damage markers and the number of symptoms were more linked to systemic inflammatory activation. This latter result reinforces the hypothesis that some patients manage to contain *P. vivax* infection in a balanced fashion, despite presenting some symptoms but without substantial tissue damage. This ‘tolerant state’ may be associated with epidemiological factors such as those reported here, confirming previously reported results [7].

Our study has some limitations. We did not have inflammatory measurements after the malaria therapy, limiting our insights about the systemic inflammation and immune restoration in our participants. Additionally, all our study participants were recruited in Brazil, and additional studies in multiple sites are needed to confirm our findings. Despite these limitations, our study adds to the current knowledge in the field by demonstrating the occurrence of disease tolerance in *P. vivax* malaria and by identifying the major determinants of this phenomenon.

## Data Availability

All data will be available in the supporting information of the manuscript once the study is accepted for publication or upon request to the corresponding author.

## Acknowledgments

The authors would like to thank Dr Luiz Marcelo Camargo (University of São Paulo) ncio and Dr Daniela Andrade for technical support in field study area; Dr Jorge Clare□ncio and Dr Daniela Andrade (FIOCRUZ) for technical help with the immunoassays, and Mr Jorge Tolentino, Ms Natali Alexandrino, Mrs Elze Leite, and Mrs Andrezza Kariny for logistic support.

## Supporting Information

**S1 File.** Raw data used in the analyses.

**S1 Table.** Characteristics of the study participants.

**S1 Figure.** Epidemiological and immune factors associated with symptomatic *Plasmodium vivax* malaria.

**S2 Figure.** Determinants of the global degree of perturbation.

**S3 Figure.** Influences of age and number of previous episodes in the inflammatory imbalance in *Plasmodium vivax* infection.

