## Supplementary Information for "Dissecting Disease Tolerance in *Plasmodium vivax* Malaria Using the Systemic Degree of Inflammatory Perturbation"

**S1 Table:** Characteristics of the study participants

| Characteristics/cytokines | Endemic Control (n=128) | Asymptomatic Malaria (n=108) | Symptomatic Malaria (n=134) | p-value |
|---|---|---|---|---|
| **Age – years** | 39 (25–50) | 44 (34–50) | 29 (19–42) | **<0.01[a]** |
| **Female sex** | 65 (50.8) | 59 (54.6) | 62 (46.3) | 0.43 |
| **Previous malaria** | 122 (95.3) | 108 (100) | 115 (85.8) | **<0.01[a]** |
| **Number of previous episodes of malaria** | 13 [9-18] | 17 [13-20] | 5 [1-11] | **<0.01** |
| **Years residing in the area** | | | | **<0.01[a]** |
| < 3yrs | 5 (3.9) | 10 (9.3) | 23 (17.2) | |
| 3-10yrs | 4 (3.1) | 29 (26.9) | 88 (65.7) | |
| >10yrs | 119 (93) | 69 (63.9) | 23 (17.2) | |
| **Haptoglobin – ng/mL** | 1.77 [0.99-1.77] | 1.72 [0.74-1.77] | 1.77 [1.26-1.77] | **0.03[a]** |
| **IFNγ– pg/mL** | 0.42 [0.22-0.71] | 0.71 [0.36-1.94] | 0.71 [0.29-4.11] | **<0.01** |
| **TNFα – pg/mL** | 0.52 [0.34-0.52] | 0.52 [0.37-0.52] | 1.91 [0.52-4.56] | **<0.01[a]** |
| **IL-10 – pg/mL** | 0.54 [0.27-0.81] | 3.92 [1.60-6.67] | 0.75 [0.60-2.48] | **<0.01[a]** |
| **IL-6 – pg/mL** | 0.55 [0.39-0.76] | 0.58 [0.34-0.91] | 2.86 [0.71-6.78] | **<0.01[a]** |
| **IL-4 – pg/mL** | 0.40 [0.19-0.66] | 0.45 [0.18-0.68] | 0.61 [0.32-2.21] | **<0.01[a]** |
| **IL-1β – pg/mL** | 0.49 [0.40-0.77] | 0.55 [0.33-0.65] | 0.57 [0.35-1.06] | 0.33 |
| **IL-8 – pg/mL** | 0.55 [0.35-0.86] | 0.92 [0.52-1.13] | 3.28 [0.78-17.85] | **<0.01[a]** |
| **IL-12p70 – pg/mL** | 0.62 [0.36-0.83] | 0.47 [0.24-0.88] | 1.28 [0.93-2.63] | **<0.01[a]** |
| **CXCL10 – pg/mL** | 0.42 [0.22-0.69] | 0.68 [0.64-0.69] | 0.63 [0.47-0.99] | **<0.01** |
| **CCL5 – μg/mL** | 0.40 [0.19-0.66] | 0.49 [0.22-0.71] | 0.66 [0.42-0.89] | **<0.01[a]** |
| **CXCL9 – ng/mL** | 0.40 [0.19-0.66] | 0.51 [0.30-0.83] | 4.79 [0.81-24.73] | **<0.01[a]** |
| **CCL2 – ng/mL** | 0.42 [0.22-0.69] | 0.44 [0.22-0.70] | 0.58 [0.28-0.69] | 0.06 |
| **AST – U/L** | 0.75 [0.29-0.92] | 1.15 [0.66-2.49] | 10.47 [3.47-46.21] | **<0.01[a]** |
| **ALT – U/L** | 0.56 [0.25-1.08] | 1.12 [0.41-1.99] | 11.7 [7.28-35.52] | **<0.01[a]** |
| **Fibrinogen – mg/dL** | 0.63 [0.46-1.10] | 0.81 [0.48-1.42] | 1.66 [0.59-3.06] | **<0.01[a]** |
| **CRP – mg/L** | 0.56 [0.35-0.88] | 0.73 [0.44-1.33] | 3.12 [0.68-7.94] | **<0.01[a]** |
| **Creatinine – mg/dL** | 0.64 [0.36-0.96] | 0.71 [0.41-1.10] | 0.71 [0.33-1.12] | 0.61 |
| **Total Bilirubin – mg/dL** | 0.82 [0.41-1.20] | 0.82 [0.46-1.44] | 1.44 [0.58-4.23] | **<0.01[a]** |
| **Direct Bilirubin – mg/dL** | 0.55 [0.34-1.03] | 1.02 [0.40-2.62] | 1.21 [0.40-3.82] | **<0.01** |
| **Indirect bilirubin – mg/dL** | 0.73 [0.35-1.13] | 1.17 [0.73-1.17] | 1.02 [0.51-3.64] | **<0.01[a]** |
| **ssMDP** | 0 [0-2.73] | 9.07 [4.19-16.38] | 48.11 [24.42-169.43] | **<0.01[a]** |

Data represent median and interquartile range or frequency (percentage). The Kruskall-Wallis test was used to compare the molecular degree of perturbation of each plasma marker between the study groups. Chi-square test was used to compare frequencies. P-values in bold font are statically significant. [a] Markers that displayed statistical significance (P<0.05) between symptomatic and asymptomatic groups.

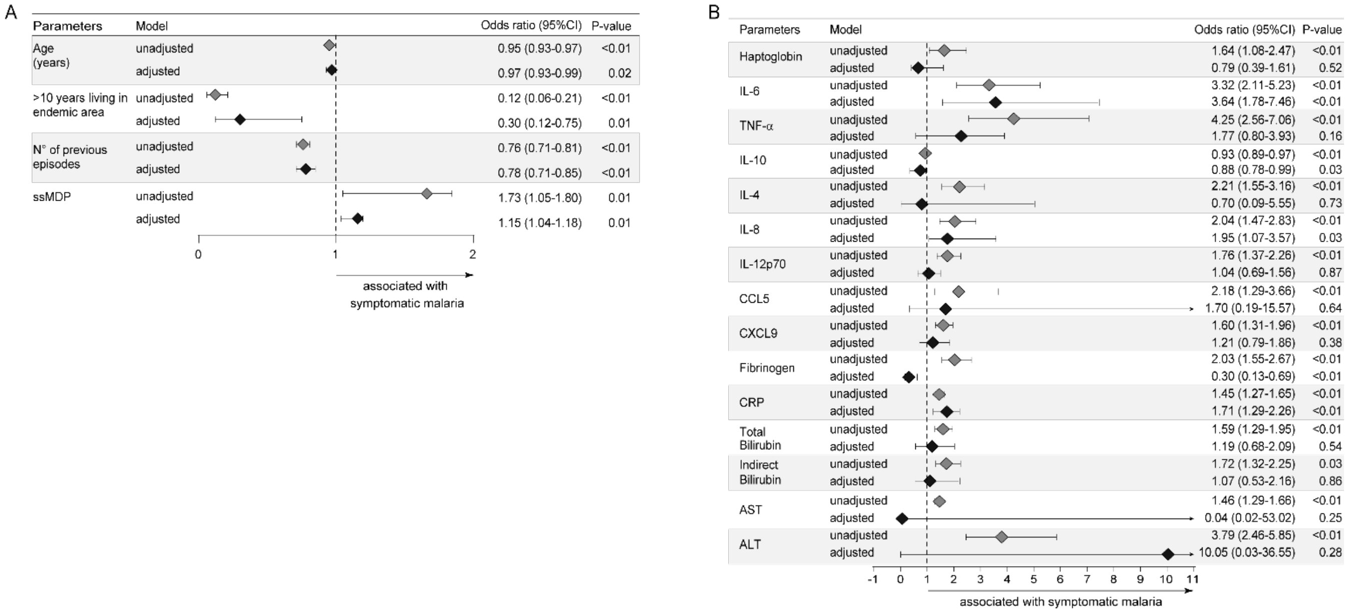


**S1 Figure: Epidemiological and immune factors associated with symptomatic *Plasmodium vivax* malaria**: Adjusted multinomial logistic regression analysis was performed with symptomatic malaria as the primary outcome. The model was composed with variables that were statistically significant ($p < 0.05$) in univariate comparisons (see univariate comparisons in Table S1).

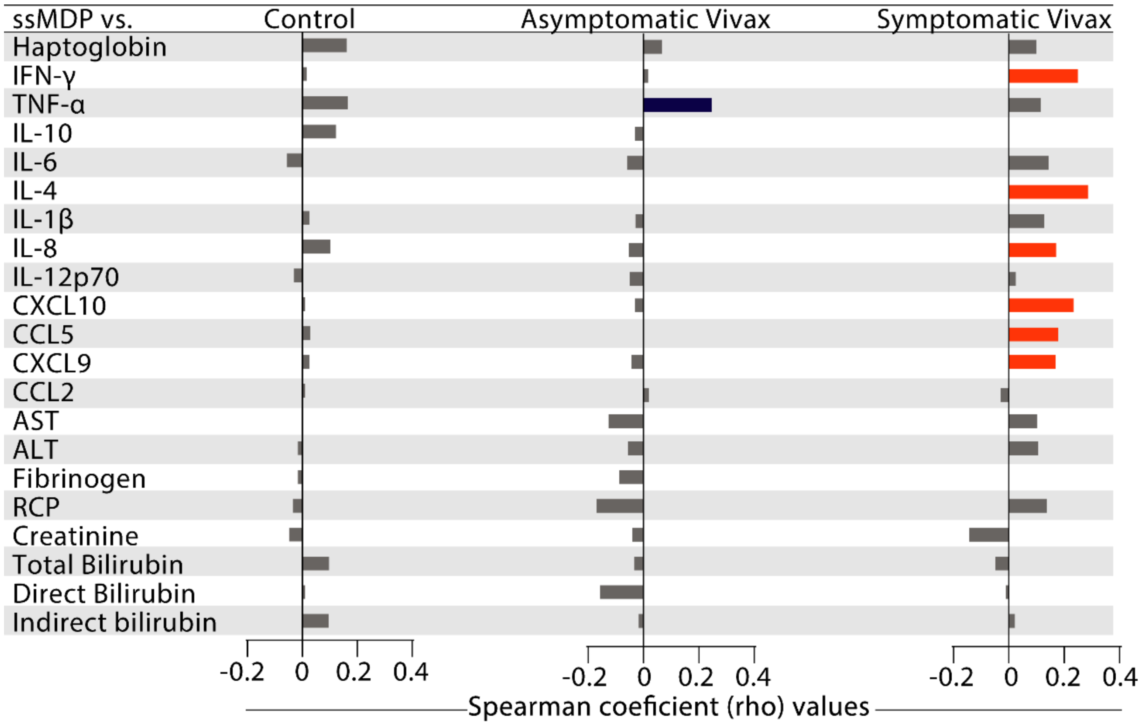


**S2 Figure: Determinants of the global degree of perturbation.** A Spearman correlation analysis was employed to identify the perturbation of each individual marker that contribute to changes in the systemic inflammatory imbalance, assessed by ssMDP score values, in each group as indicated. Colored bars infer the correlation with P-value<0.05 after adjustment for multiple comparisons. Abbreviations (alphabetic order): ALT: alanine aminotransferase; AST: aspartate aminotransferase; CCL: C-C motif chemokine ligand; CXCL: C-X-C motif chemokine ligand; CRP: C-reactive protein; IFN: interferon; IL: interleukin; TNF: tumor necrosis factor.

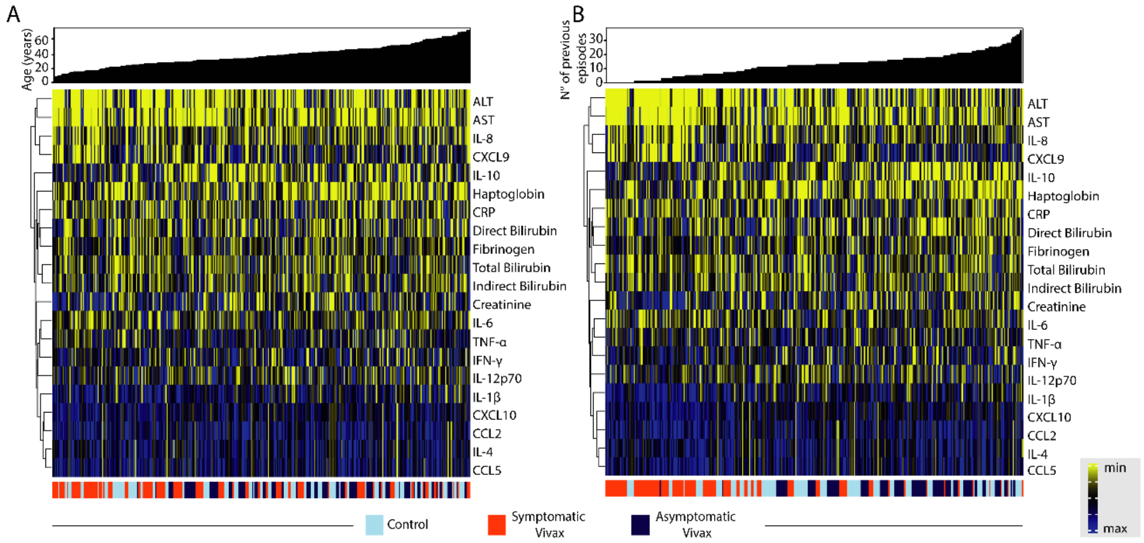


**S3 Figure: Influences of age and number of previous episodes in the inflammatory imbalance in *Plasmodium vivax* infection.** Hierarchical cluster analysis of Log-10 transformed and z-score normalized using Ward's method with 100X bootstrap was employed to depict the overall perturbation of inflammatory and biochemical markers in study population. The participants were grouped based on age (A) and number of previous Malaria episodes (B).
